# Associations between RetNet gene polymorphisms and efficacy of orthokeratology for myopia control : sample from a clinical retrospective study

**DOI:** 10.1101/2024.09.18.24313851

**Authors:** Ruijing Xia, Xiangyi Yu, Lulu Peng, Zhenlin Du, Xiaoguang Yu, Shilai Xing, Fan Lu, Xinjie Mao

## Abstract

**Background:** To study how clinical and genetic factors control the effectiveness of orthokeratology lenses in myopia.

**Methods:** In this study, we conducted a retrospective clinical study of 545 children aged 8–12 years with myopia who were wearing orthokeratology lenses for one year and performed whole-genome sequencing (WGS) for 60 participants in two groups, one with rapid axial length progression of larger than 0.33 mm and the other with slow axial length progression of less than 0.09 mm. Genes in the RetNet database were used to screen candidate genes that may contribute to the effectiveness of orthokeratology lenses in controlling myopia.

**Results:** We found that children with a greater baseline eye axial length, greater spherical equivalent (SE) and greater age had better myopia control with orthokeratology. We observed a significant excess of nonsynonymous variants among those with slow myopia progression, which were prominently enriched in retinal disease related genes. We subsequently identified *RIMS2* (OR=0.01, *p*=0.0075) and *LCA5* (OR=6.96, *p*=0.0080) harboring an excess number of nonsynonymous variants in patients with slow progression of high myopia. Two intronic common variants rs36006402 in *SLC7A14* and rs2285814 in *CLUAP*1 were strongly associated with axial length growth. Together, our finding identified novel genes associated with the effectiveness of orthokeratology lenses therapy in myopic children and provide insight into the genetic mechanism of orthokeratology treatment.

**Conclusion:** The effectiveness of orthokeratology lenses treatment involved interindividual variability in controlling axial length growth in myopic eyes. The efficacy increased when patients carried more nonsynonymous variants in retinal disease-related gene sets. Our data will serve as a well-founded reference for genetic counseling and better management of patients who choose orthokeratology lenses to control myopia.

## Background

Refractive error, which accounts for the largest percentage of visual impairments, is a loss of vision due to a change in the shape of the eye that prevents light from being accurately refracted and focused on the retina. Owing to its increasing prevalence, myopia has become a global public health problem. Globally, 10% to 30% of adults suffer from myopia, and in Western populations of the United States and Europe, the prevalence of myopia is approximately 40% to 50% in young adults, with an even higher probability of 80% to 90% in some countries in East and Southeast Asia^[1][2][3][4][5][6][7][8]^. Myopia is also strongly associated with a number of ocular diseases, such as cataract, glaucoma, and myopic macular degeneration^[9]^.

Myopia can usually be corrected by spectacles, contact lenses or refractive surgery to provide good vision. There are many ways to control the progression of myopia^[10]^. Atropine eye drops, orthokeratology, and peripheral defocus modifying contact lenses or spectacles are more effective at controlling axial length elongation^[11]^. Orthokeratology lenses focuses light in front of the peripheral retina primarily by changing the curvature of the cornea which returns to its original way, thereby refocusing the image centrally at the fovea^[12]^. This causes the image contour to focus on the fovea in the center and the macula in the periphery, resulting in myopic defocus, which slows the progression of myopia.

However, there are strong individual differences in the control effects of orthokeratology lenses. Some have fairly good control, while some have very limited or even accelerated control^[13][14][15][16]^. In several studies, the baseline corneal stiffness, lower baseline myopia, younger initial age and higher parental myopia have been identified as factors influencing the effectiveness of orthokeratology lenses control^[17]^ ^[18]^. Although many risk factors for the efficacy of orthokeratology lenses for myopia control have been revealed, the genetic factors underlying orthokeratology lenses treatment are still unknown.

This study aimed to explore the genetic characteristics of 289 retinal disease-related genes from the Retinal Information Network database (RetNet, https://retnet.org/) and clinical features in a cohort of orthokeratology lenses users. We also investigated whether these genes and specific SNPs are associated with the effectiveness of orthokeratology lenses treatment. Through this research, we hope to uncover potential relationships between genes related to retinal function and the effectiveness of orthokeratology lenses treatment, thereby enhancing our understanding of the genetic background of myopia treatment and providing new insights for developing personalized vision correction.

## Methods

### Subjects

This 1-year study used a prospective and randomized eye crossover study design. The protocol and documentation conducted in this study received full approval from institutional ethics committee of Wenzhou Eye and Vision Hospital with approval number 2023-059-K-48-05.

Written informed consent was obtained from each participant. The orthokeratology lenses used in this study included four-zoned reverse-geometry lenses (Eulcid, USA and Lucid, Korea)^[19]^. Each subject underwent comprehensive baseline eye examination, including slit-lamp examination, testing for spherical equivalent (SE) of refraction, uncorrected visual acuity and best-corrected visual acuity, and axial length (AL) via Zeiss IOL-Master 500. All children were treated by doctors who had worked in the field of orthokeratology lenses at the hospital for more than 10 years. The doctor ordered the best lens for the subject based on the basis of each subject’s corneal topography and evaluated the fit of the evaluation of corneal fluorescein pattern analysis. A total of 1,538 myopic patients who were initially wearing orthokeratology lenses were enrolled in the study. These 1,538 samples were critically reviewed and screened, leaving 545 samples with complete data and meeting the filter criteria. Each sample contained clinical phenotypic data for both eyes. The amount of AL growth in both eyes of each sample was considered as an indication of the effectiveness of orthokeratology lenses in controlling myopia. We counted the amount of AL growth per year in each sample and took the anterior and posterior quartiles of AL growth as the case group (well controlled) and control group (poorly controlled, Figure 1b). In total, there were 100 cases with annualized AL growth less than or equal to 0.09mm, and 100 controls with annualized AL growth greater than or equal to 0.33mm. We randomly selected 30 cases and 30 controls for whole-genome sequencing with their written informed consent as the final analysis cohort. Figure 1a shows the entire study outline.

**Figure 1.**
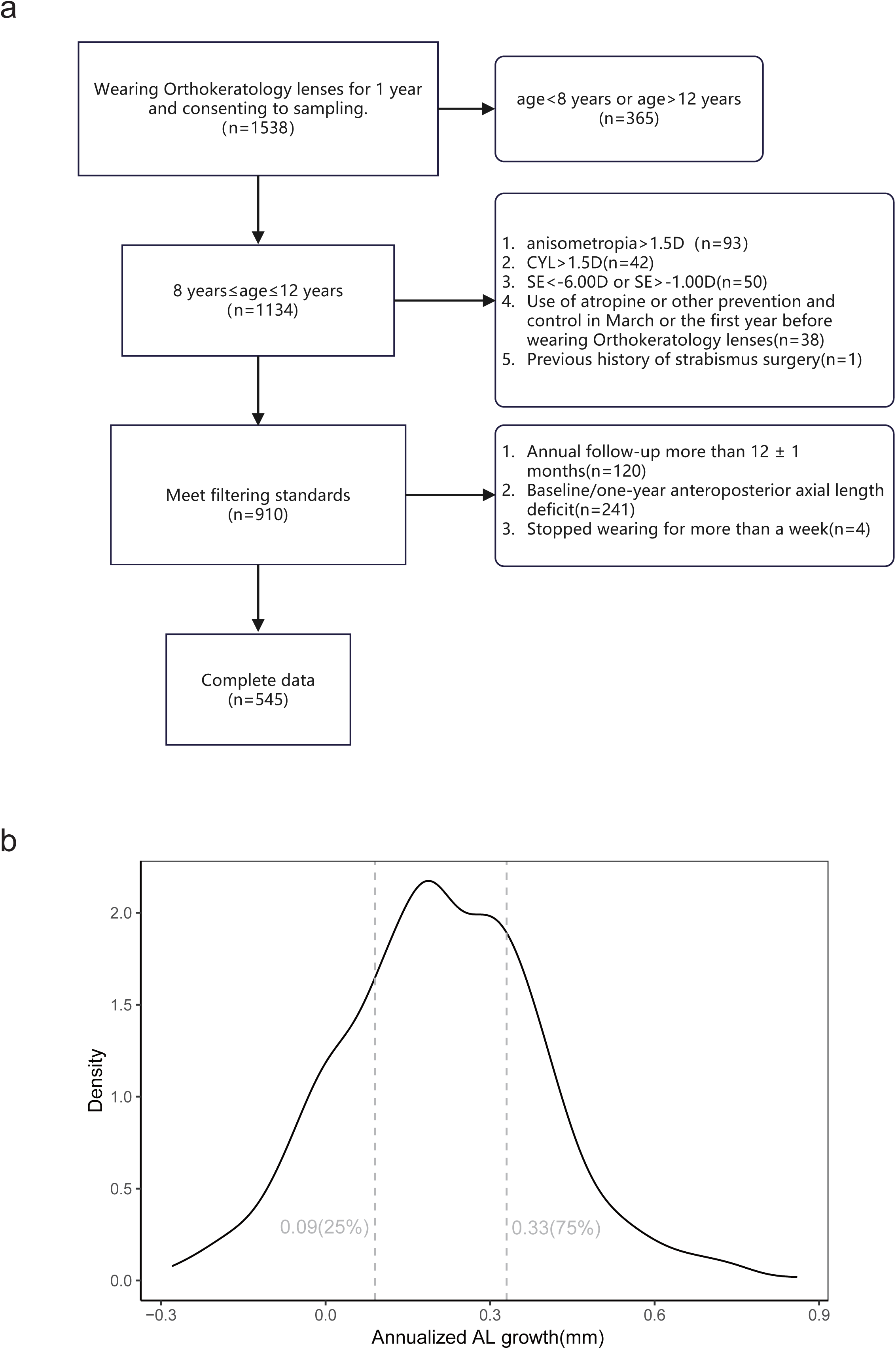
Study flowchart and sample screening process. (a). screening process for 1538 study participants. (b). selecting populations with distinct control effects based on quartiles.

### Sequencing and variant calling

The genomic DNA of all subjects was isolated from oral swabs via standard procedures. Whole-genome sequencing was performed using DNBSEQ-T7. Variant detection and joint genotype calling analyses were conducted based on the Sentieon DNAscope pipeline (Sentieon Inc., version 202308)^[20]^. The sequence reads of each sample in FASTQ format were aligned against the human reference genome (GRCh38) using BWA-MEM^[21]^. The alignment file was sorted using Sentieon sort algorithm, and the Sentieon Dedup algorithm was used to mark duplicated reads. Then SNVs and indels were called in genomic variant call format (GVCF) using Haplotyper. The Sentieon GVCFtyper jointly called subjects as a cohort.

### Quality control

We applied standard variant-level quality controls. We excluded variants for further analysis if the had an average genotype depth (DP) < 20 and a genotype quality (GQ) < 40. We detected population outliers and stratification using a method based on principal component analysis, which indicated that the affected individuals and control subjects were genetically matched for all sequenced samples using Plink 2.0^[22]^(Additional File 1). PCs 1 to 10 were assessed for their associations with disease phenotype status using a generalized linear model (GLM) and then included in the GWAS as covariates. A population check was conducted on east Asian populations including CHB (China Beijing), CHS (China South), CDX (China Dai), JPT (Japan) and KHV (Korean) individuals from the 1000 Genome Project (1KG)^[23]^.

### Variant Annotation

The annotation of variants was performed with Ensembl’s Variant Effect Predictor (VEP v.0.1.16) ^[24]^for the human genome assembly GRCh38. We used population allele frequency (AF) data from the following databases: 1000 Genomes, ESP and Genome Aggregation Database (gnomAD)^[25]^. We employed multiple in silico prediction algorithms, including PolyPhen-2^[26]^, SIFT^[27]^, Combined Annotation Dependent Depletion (CADD)^[28]^, LOFTEE and SpliceAI^[29]^ plugins to generate additional bioinformatic predictions of variant deleteriousness. Protein-coding variants were annotated into the following three classes: (1) synonymous; (2) nonsynonymous (3) noncoding. In detail, nonsynonymous variants were classified as PTV variants and missensen varinat including following annotation terms:(1)PTV:“frameshift_variant”, “splice_acceptor_variant”, “splice_donor_variant”, “stop_gained”, “start_lost”, “stop_lost”, protein_altering_variantor SpliceAI > 0.5 & LoF=HC. (2) Missense: inframe_insertion, “iqnframe_deletion”, “missense_variant”. The synonymous variants was predicted as “synonymous_variant”. Finally, non-coding was classified as “intron_variant”,“intergenic_variant”, “downstream_gene_variant”,“3_prime_UTR_variant”, “5_prime_UTR_variant”,“mature_miRNA_variant”,“regulatory_region_v ariant”,”non_coding_transcript_exon_variant” and “upstream_gene_variant”.

### Gene-set burden analysis

To estimate the extent to which variants with different allele frequencies and different functions are over-represented in individuals with different control effects, we conducted burden tests across the entire genome and 289 RetNet genes^[30]^. Common and rare variants were differentiated according to allele frequency from ChinaMap^[31]^, with minor allele frequency (MAF) less than 0.05 as rare variants and vice versa for common variants. For RetNet genes, we performed a logistic test by regressing the case-control status on certain classes of variants aggregated across target gene set in an individual and adjusting for sex, the baseline AL, the top ten PCs, and the genome-wide variant count.

### Gene-based collapsing analysis

For the gene-based test, we restricted our testing to common variants annotated as nonsynonymous. To assess whether a specific gene exhibited an over-representation or under-representation of common nonsynonymous cases, we performed five gene-level association tests, including Fisher’s exact test, logistic, SKAT^[32]^, SKAT-O^[33]^ and Magma^[34]^, with previously defined covariates (sample sex, PC1-PC10).

### Cell type enrichment

We acquired a single-cell RNA-seq (scRNA-seq) expression matrix and metadata of developing human embryonic eyes from the Broad Institute Single Cell Portal (https://singlecell.broadinstitute.org/, SCP1311) and performed a scRNA-seq analysis in R4.3.2.

### Single-variant association analysis

We estimated associations of common variants (MAF > 0.05) by using Saige^[35]^, fastGWAS^[36]^, PLINK, MLMA-LOCO ^[37]^and EMMAX^[38]^ tests and corrected for the first ten PCs.

### Statistical analysis

Statistical analyses were performed using R4.3.2. The differences in phenotype and sequencing quality between groups were compared by Student’s t-test. These phenotypes were also evaluated for phenotype-genotype correlation by Wilcoxon rank sum test. Statistical significance was defined as a *P*-value less than 0.01. ∗ *P* < 0.05, ∗ ∗ *P* < 0.01, ∗ ∗ ∗ *P* < 0.001.

## Results

### Subject Demographics

After a rigorous data review, 545 of 1538 patients were considered complete and compliant. At baseline, their age ranged from 8 to 12 years(10.12±1.27), their SE refractive errors ranged from −1 to −6 (OD: −3.11 ± 1.08; OS: −3.08 ± 1.09) diopters (D), and their AL ranged from 22.96 to 27.94 (OD: 24.85 ± 0.80; OS: 24.83 ± 0.80)mm. The orthokeratology lenses used in this study has no difference in the amount of annualized AL growth(Eulcid, USA and Lucid, Korea. P=0.66, Wilcoxon rank sum test). A total of 100 cases and 100 controls were selected on the basis of quartiles of annualized AL growth (Methods), and accordingly 30 cases and 30 controls were randomly selected for sequencing and genetic analysis.

### Clinical influences on the effectiveness of OK lens control

After screening 545 samples of acceptable quality, we focused on the correlation between the baseline phenotype (age,SE,AL) and annualized AL growth in these samples. We found that all three phenotypes correlated with the AL growth in both eyes (Figure 2). The older samples had smaller AL growth (OD: cor = −0.26, *p* = 1.3e-09; OS: cor = −0.23, *p* = 3.5e-08). Similarly, the same negative correlation was found in the statistics for baseline AL (OD: cor = −0.2, *p* = 1.9e-06; OS: cor = −0.24, *p* = 1.3e-08) and SE (OD: cor = −0.24, *p* = 1.9e-08; OS: cor = −0.29, *p* = 7.6e-12).

**Figure 2.**
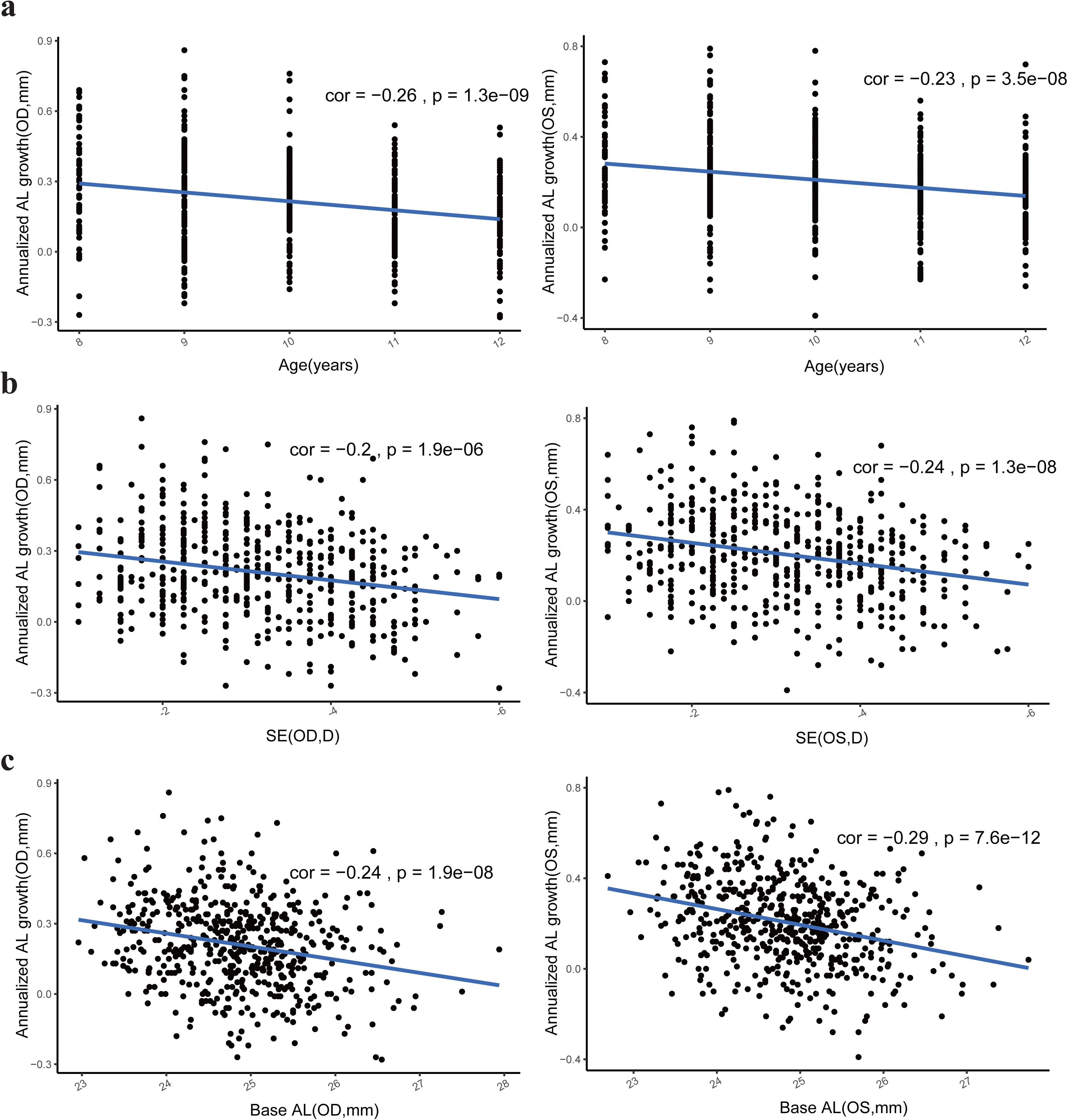
Correlation between Baseline Data and annualized AL growth. The correlation between AL growth and (a) age, (b) SE, (c) baseline AL.

Furthermore, we analyzed the baseline phenotypes of 30 samples with well controlled AL growth and 30 samples with poorly controlled AL growth. Significant differences in the amount of AL growth were found between the two groups (OD: *p*=2.11e-24; OS: *p*=2.29e-46. Figure 3a). There were no significant differences in age distribution or SE between two eyes (Figure 3b,c). In contrast, a significant difference in baseline AL was found between the two groups (OD: *p*=0.00313; OS: *p*=0.00608. Figure 3d).

**Figure 3.**
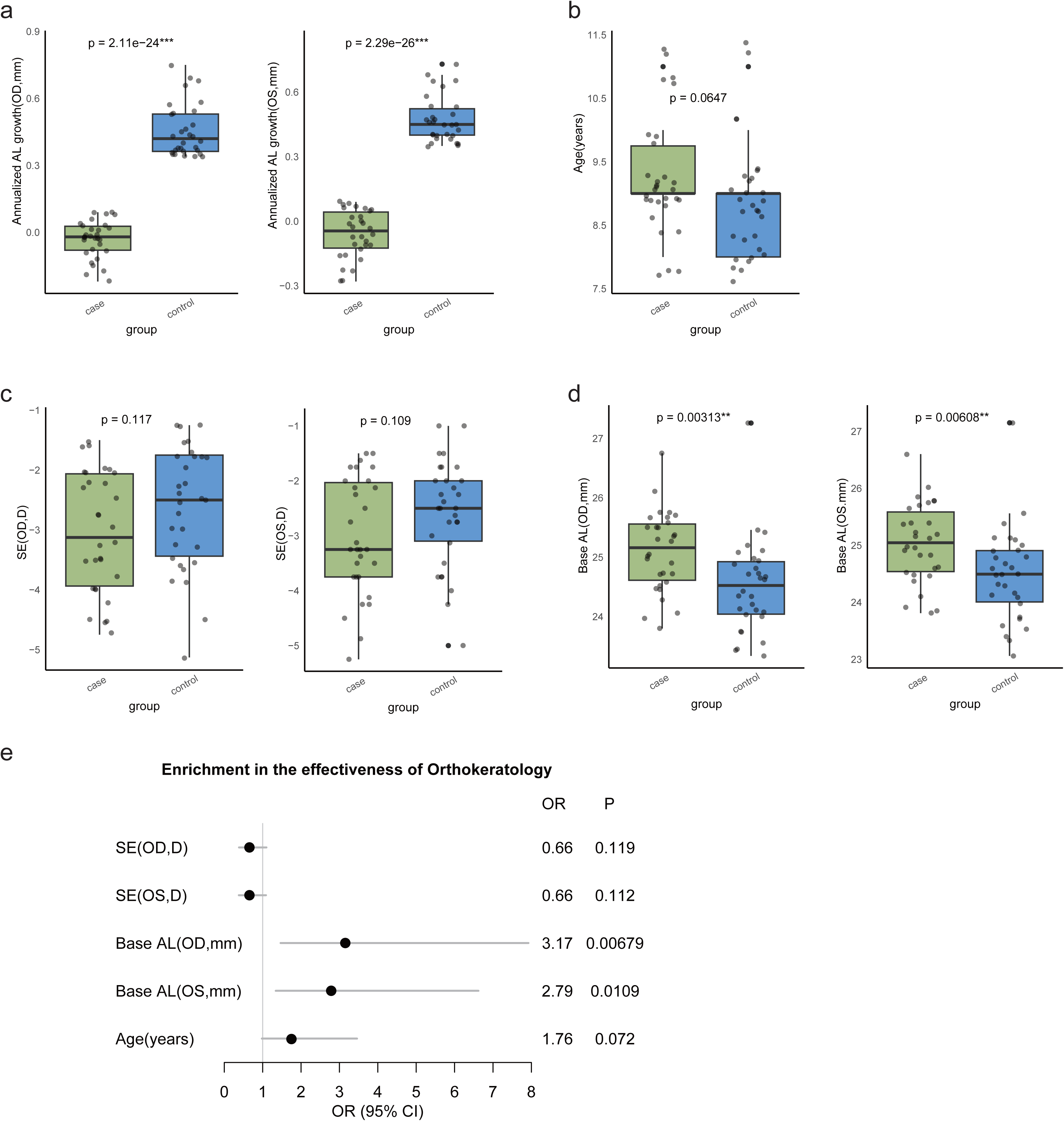
Differences in Baseline Data and Clinical Factors Affecting Control Effectiveness. Difference between case and control in (a) AL growth, (b) age, (c) base SE and (d) base AL. (e) logistic regression in the effect factor of orthokeratology lenses control.

The effects of different baseline phenotypes on orthokeratology lenses in myopia control were subsequently tested by using a logistic regression model (Figure 3e). We identified a significant enrichment of baseline AL among cases with a positive effect compared with controls (OD: OR=3.17, *p*=0.00679).

### Whole-genome sequencing of 60 samples

After stringent quality control, we used WGS data from 30 cases and 30 controls in the discovery stage. A total of 7,644,581 bialleic variants were used for further analysis, including 3,233,468 common variants and 3,254,265 rare variants divided by ChinaMap. Sequencing quality was not significantly different between the cases and controls (Additional File 1). These remaining samples were all ancestry-matched, closely resembling CHB and CHS ancestry in the 1000G genome.

### Excesses of gene-set based nonsynonymous variants

To aggregate multiple alleles of presumed similar impact in retina disease-associated genes, we adopted a complementary strategy, focusing on variants in different functional regions, and sought to improve the ability to detect variant associations by exploiting the more robust functional annotation of coding variation. We first evaluated the associations among the burden of all variants, common variants and rare variants by a firth-logistic models, and then dissected the burden test into the RetNet gene set. Specifically, the model used Firth-based logistic regression and incorporated sample sex, principal component (PC) 1-10, the total genome count and sample base AL. There was no significant difference in any type of variants between cases and controls when the gene-set was not restricted (Additional File 2).

Gene sets covering different biological processes and pre-experimental validations could refine our understanding of the mechanisms of associations between the variants and the control effects of orthokeratology lenses and help to derive potential biological hypotheses for subsequent detailed analyses. We chose the RetNet gene collection to explore the associations between retina-associated biological pathway genes and orthokeratology lenses control effects. After restricting the gene set, we observed significant enrichment of nonsynonymous variants in cases (OR=1.34, *p*=0.00106, Figure 4a).

**Figure 4.**
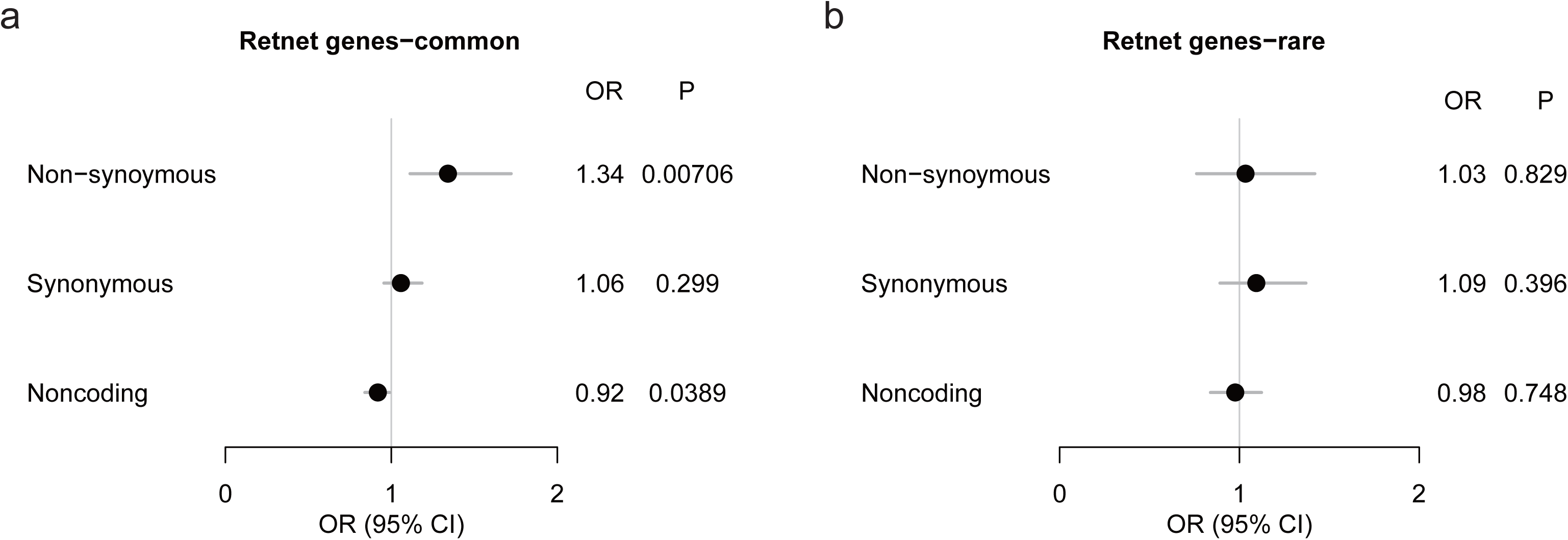
Gene-set polygenic burden test of different type of variant. Gene-set burden analysis in nonsynonymous, synonymous and noncoding variants with a MAF (a) >0.05, (b) <0.05.

### Gene-based common variant association analysis

To identify genes associated with the effect of orthokeratology lenses on myopia control, we performed an association analysis in which individuals were categorized on the basis of presence or absence of common nonsynonymous variants. The genes significantly associated with positive effects included 3 variants in *RIMS2* (OR=0.01, *p*=0.0075) and *LCA5* (OR=6.96, *p*=0.0080; Figure 3a, b. Table 1). Cell-type specificity analysis of data from whole eyes consistently revealed that *RIMS2* was mainly expressed in retina and that *LCA5* was expressed at low levels in various eye tissues (Figure 5c, d, e). Furthermore, when the tissue was restricted as retina, both genes presented the strongest expression in rod cells (Figure 5f, g).

**Figure 5.**
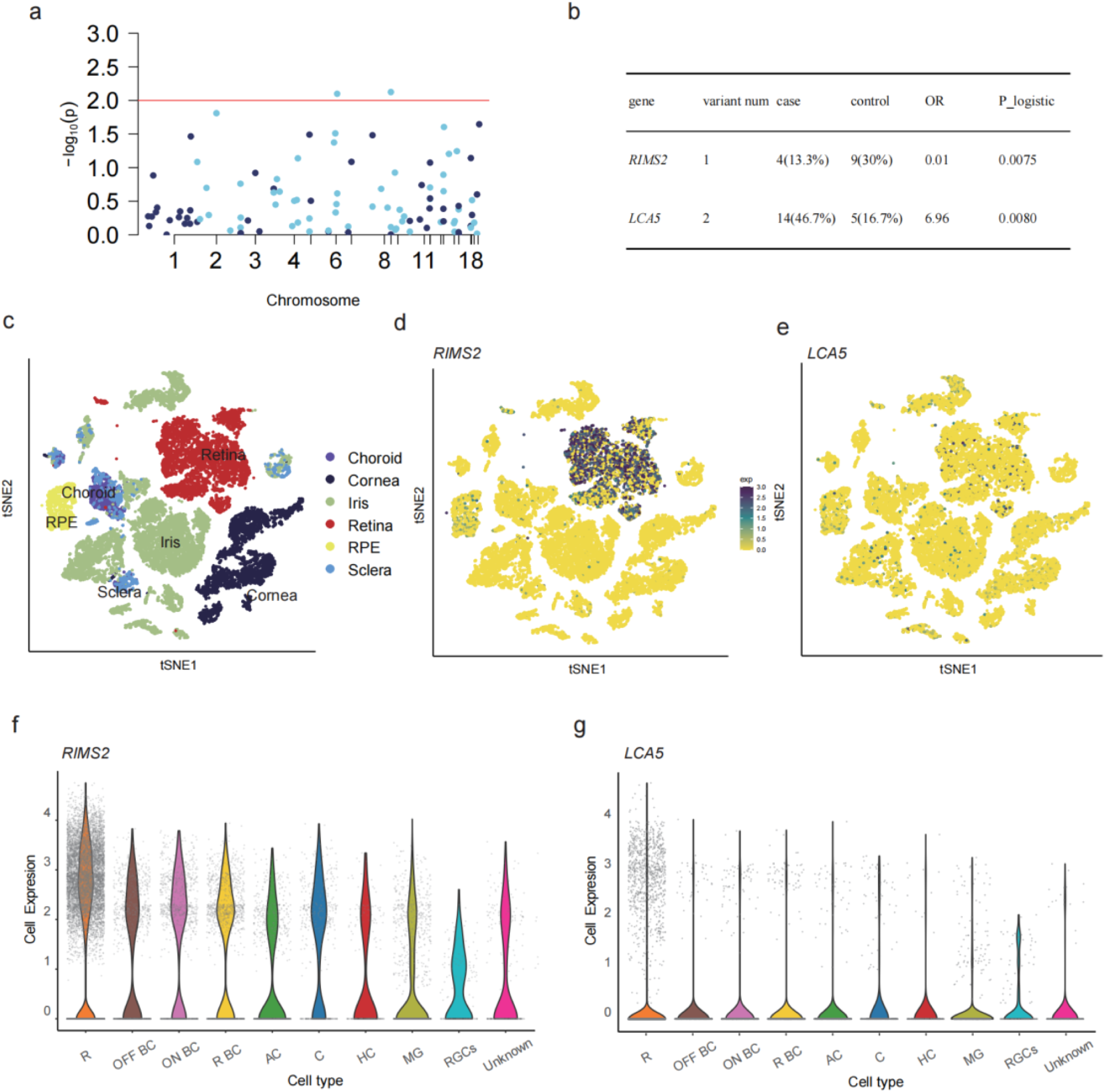
Collapsing analysis identifies 2 genes affect the efficacy of orthokeratology. (a), Manhattan plots of the gene-based collapsing analysis. An excess of nonsynonymos within RetNet genes were tested using Logistic regression, Red line, *P* = 0.01. (b), 2 genes from collapsing analyses under the same model are shown, including the exact numbers of all qualifying cases and controls and statistical calculations of association (OR and *P*). tSNE of (c) all tissues single-cell data with cell colored based on the expression of (d) *RIMS2* genes and (e) *LCA5*. Gene expression levels are indicated by shades of blue. Violin plot of expression in sub celltype of retina in (f) *RIMS2* and (g) *LCA5.* Abbreviations are as follows: R: rod cells, OFF BC: OFF bipolar cells, ON BC: ON bipolar cells, R BC: Rod bipolar cells, AC: Amacrine cells, C: Cone, HC: Horizontal cells, MG: Muller glia.

**Table 1.** All qualified variants in *RIMS2* and *LCA5* in 30 cases and 30 controls with ChinaMap MAF > 0.5%.

| Chromosome | Position | SNP | HGVS.c | HGVS.p | Variant type | PolyPhen | MAF in ChinaMap | Alleles in 30 cases | Alleles in 30 controls |
| --- | --- | --- | --- | --- | --- | --- | --- | --- | --- |
| *RIMS2* |  |  |  |  |  |  |  |  |  |
| chr8 | 104093618 | rs55788818 | c.3841C>T | p.Arg1281Cys | missense variant | probably_damaging(0.998) | 0.15 | 4 | 9 |
| *LCA5* |  |  |  |  |  |  |  |  |  |
| chr6 | 79487131 | rs1875845 | c.1967G>A | p.Gly656Asp | missense variant | benign(0) | 0.25 | 15 | 6 |
| chr6 | 7951881 | rs34068461 | c.77A>C | p.Asp26Ala | missense variant | possibly_damaging(0.808) | 0.22 | 14 | 5 |

### Single-variant association analyses in RetNet genes

We examined all common variants of RetNet genes that passed standard quality control for associations test, utilizing a generalized mixed-based method (SAIGE) capable of accommodating population structure, a sparse genetic relationship matrix and relatedness. The discovery analysis identified variants that reached the significance level including 16 SNPs (Table 2).

**Table 2.**
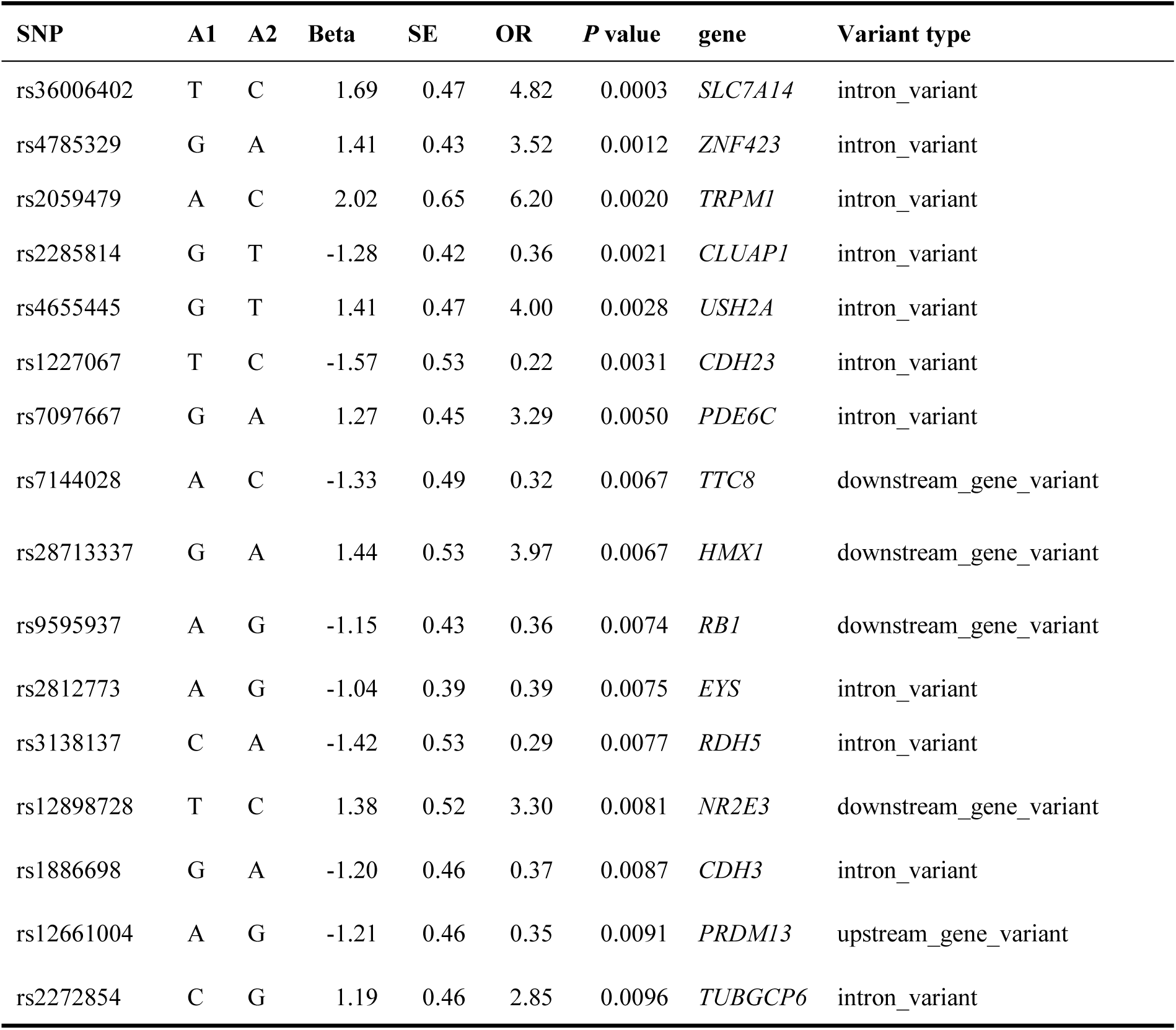
The most significant single-variant associations for orthokeratology lenses effect identified by SAIGE analysis.

We further explored the relationships between these 16 variants and the axis growth. We found that, compared to those with the wild type, homozygous carriers of rs36006402 have a lower AL growth(*p*=0.005). Greater axial growth was found in both homozygous (*p*=0.0084) and heterozygous (*p*=0.0096) carriers of rs2285814 (Figure 6).

**Figure 6.**
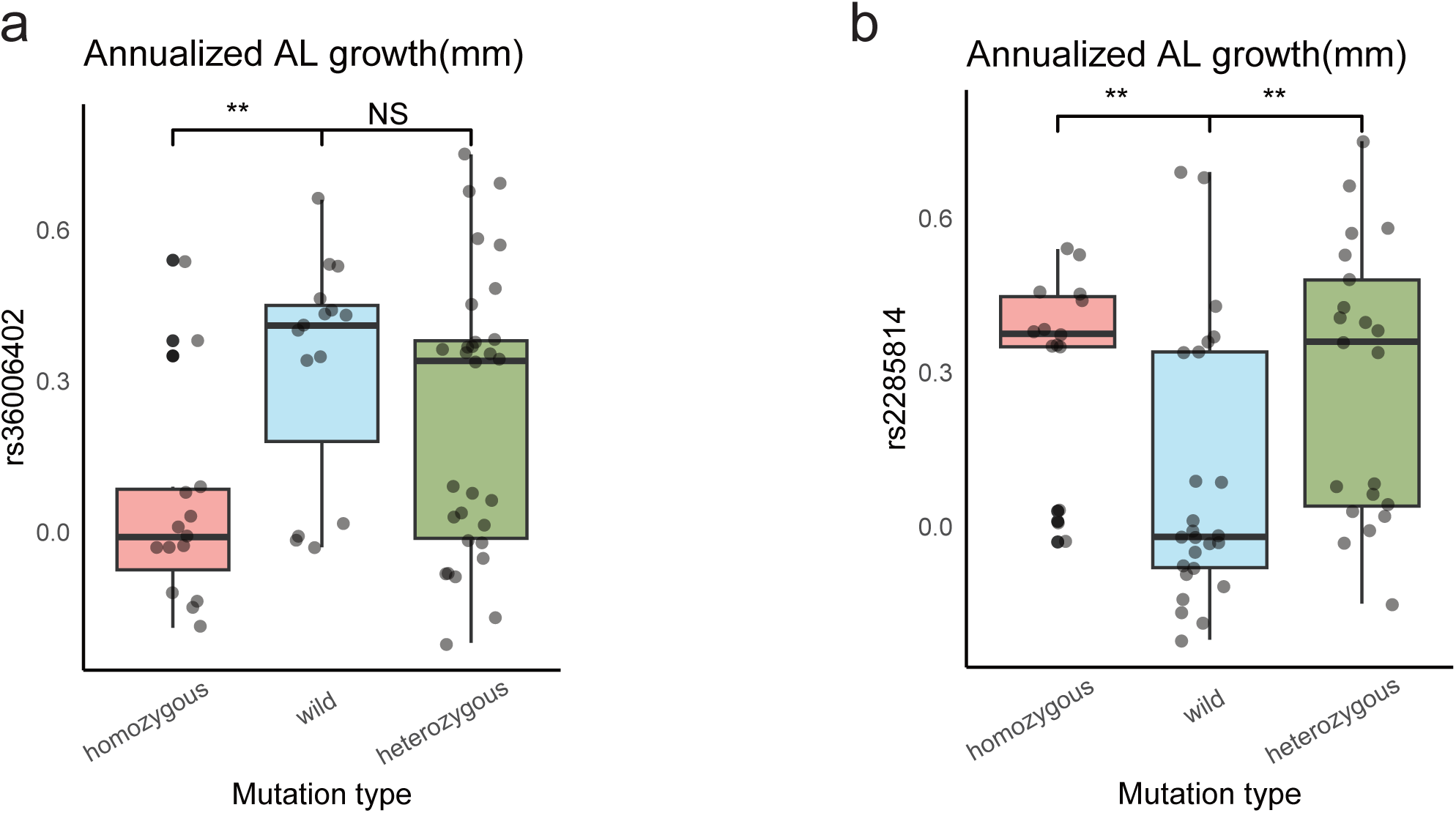
Cumulative difference of AL growth. Differences in annual AL growth between populations carrying heterozygous, homozygous, and wild-type variants of (a) rs36006402 and (b) rs2285814.

## Discussion

In the present study, we compiled one of the largest orthokeratology lenses cohorts genotyped via whole-genome sequencing. With this comprehensive dataset, we explored not only the clinical factors that influence the effectiveness of orthokeratology lenses in controlling myopia through the annual increase in AL, but also the genetic landscape and underlying biological mechanisms of the efficiency of orthokeratology.

Through rigorous and comprehensive data screening, 545 samples were retained out of a amount of 1583 samples. The baseline data revealed a significant negative correlation between age, initial myopia progress and the amount of ocular AL growth. Younger individuals with lower baseline AL and SE experienced greater AL growth.

Age has been found to be negatively correlated with AL growth after wearing orthokeratology lenses in previous studies^[8][17][39][40][41][42]^. This may be because that the eye axis naturally decreases with age within the children population^[43][44][45]^. There is a relatively high risk of progression to high myopia among children with early school-age myopia onset^[46]^. Moreover, younger myopic patients harbor more variants in genes that affect vision^[47][48][49]^. These findings indicated the influence of genetic factors on the progression of myopia. Several studies have also reported an association between higher baseline SE and lower amounts of ocular axis growth^[50][51][18][15]^.Similarly, the conclusion that a longer baseline AL can retard axial growth while wearing orthokeratology lenses has been confirmed by multiple studies^[52][53]^. A natural slowing of AL growth may occur once the eye approaches a specific threshold for myopia and AL. These findings also suggest that the use of orthokeratology lenses in older children with greater degreess of myopia may be more effective in slowing the progression of AL growth. On the other hand, for younger children with lower degreess of myopia, a combination of treatment methods, such as low-concentration 0.01% atropine^[54][55]^ ^[56]^, may be needed for optimal results as previous research has shown that combined use is effective in younger children^[45]^.

Through the quartile division, we identified 100 patients whose annual AL growth was less than 0.09 and 100 patients with annual AL growth more than 0.33. This situation specifies that there is a large difference in the degrees of acceptance of OK lenses by each individual. We further selected 30 cases from each group for whole-genome sequencing. There was a very significant difference in the amount of AL growth between the two groups. Differences in the baseline AL were found between the two groups. Additionally, baseline AL was also considered a factor that promoted the efficacy of orthokeratology lenses according to the logistic regression analysis. This conclusion further validates the findings from the correlation analysis mentioned before that a more severe initial state of myopia might be a potential factor for achieving better outcomes with the subsequent use of orthokeratology lenses. These findings suggest that the baseline AL can be used as a factor for assessing and predicting the effectiveness of myopia control in patients.

Subsequently, we explored the genetic characteristics of the samples that influence the effectiveness of orthokeratology lenses at the genetic level by a whole-genome sequencing. Gene-set burden analysis of the RetNet gene set revealed that common nonsynonymous variants promote the effectiveness of orthokeratology. Hyperopic shift occurs in central retina and myopic defocus in the peripheral retina after overnight orthokeratology^[14][57][58]^which suggests an inextricable role for retinal photoreceptor mechanisms in orthokeratology lenses wear. Visual signals from the peripheral retina have a strong influence on eye growth^[59][60]^, and biological processes mediated by genetic variants in the RetNet gene set may affect the defocusing effect of orthokeratology^[61]^, thereby enhancing or preventing the inhibitory effect on AL growth produced by wearing orthokeratology. In addition, mutations in genes in RetNet have been implicated as part of the evidence for early-onset high myopia^[62][63][64]^. The myopic pathway caused by variants in the RetNet gene may also lead to more pathologically early-onset myopia than those in late-onset^[62]^ myopia, such that younger patients have even higher annualized AL growth.

Further after that, we found that two genes were associated with orthokeratology lenses control. *RIMS2* plays a negative role in the control effect of orthokeratology. *RIMS2* has the highest expression in the retina among all ocular tissues. The maximum expression was detected in rod cells in the retina. In contrast, nonsynonymous variants of *LC5A* facilitated the validity of orthokeratology. The expression of *LCA5* was the same highest in the Rod cells. *RIMS2* is the primary large *RIM* isoform found at photoreceptor ribbon synapses, and is crucial for maintaining normal synaptic connections. Mutations in *RIMS2* may result in post-photoreceptor defects affecting both the cone and rod signaling pathways^[65]^,foveal changes and inner retinal thinning^[66]^. In this study, *RIMS2* may influence the effectiveness of orthokeratology lenses by affecting the hyperopic shift in the retina, potentially through its role in synaptic neurotransmitter transmission in Rod cells. The *LCA5* gene is associated with Leber congenital amaurosis (LCA), a hereditary retinal disease that severely affects vision. Mutations in the *LCA5* gene can lead to functional impairment and structural abnormalities in the retinal photoreceptor cells^[67][68][69]^. However, no phenotypic differences were detected between samples harboring these two gene variants and those without. Their age, SE, and baseline AL remained consistent. This might be due to the limitation of the sample size. In the future, larger-scale data is needed to explore the relationships between phenotypes and molecular characteristics at the genetic level, in order to uncover the underlying control mechanisms.

Subsequently, at the SNP level, association analysis revealed 16 mutations located in 16 different genes related to the effectiveness of orthokeratology. These signals suggest that the effectiveness of orthokeratology lenses is linked to certain genetic characteristics. Among these 16 variants, rs36006402 and rs2285814 were found to be significantly associated with the AL growth. Individuals carrying the homozygous rs36006402 variant showed decreased AL growth compared with those with the wild-type or heterozygous variant, whereas those carrying rs2285814 in both the homozygous and hetozygous manners had increased AL growth. The rs36006402 (OR=4.8, *p*=0.0003) is located in the intron area of the *SLC7A14* gene and is inherited in a recessive manner. *SLC7A14* plays an important role in retinal development and visual function^[70]^. The rs2285814 (OR*=*0.36*, p*=0.0021) also occurs at an intron position in the *CLUAP*1 gene. CLUAP1 is associated with the intraflagellar transport (IFT) complex B group of proteins and undergoes IFT in both invertebrates and vertebrates, which has been associated with photoreceptor maintenance^[71][72]^. After all, further analysis is required to do the replication and functional validation of these associations.

Our study has several strengths. First, to date, no research has explored the genetic associations of the effectiveness of orthokeratology, with studies only addressing only differences in clinical information. This study employs a genome wide sequencing strategy in the Chinese population, which provides high-density coverage of noncoding regions, offering an opportunity to identify novel susceptibility loci. The unveiling of the first genetic study on orthokeratology lenses effectiveness cohort highlights a significant milestone in the field, offering a wealth of insights into the genetic underpinnings and clinical manifestations of these methods for myopia control with complex effects.

The limitations of this work primarily include the small sample size for GWAS analysis, which leads to insufficient statistical power and biased effect size estimation. Large-scale studies enable the confident identification of variants with small effect sizes and low allele frequencies, thereby facilitating a deeper understanding of the genetic basis of orthokeratology lenses effectiveness. Accordingly, these findings warrant replication in additional cases to further investigate the broader impact of these genes on the effectiveness of orthokeratology.

## Conclusion

In summary, our study revealed that age, baseline AL, and baseline SE are clinical factors affecting the effectiveness of orthokeratology. We initially conducted a WGS-based association study restricted to a retinal disorder gene set in a sizable Chinese cohort, which not only enhances the efficiency of array-based genetic studies for identifying both common and low-frequency susceptibility variants but also helps depict the genetic etiology of orthokeratology lenses effectiveness. The successful pursuit of subsequent steps will refine current heuristics for the clinical decision-making of this complex treatment method.

## Supporting information

Addditional File 1

Addditional File 2

## Data Availability

All data produced in the present study are available upon reasonable request to the authors

## Abbreviations

(WGS): Whole-genome Sequencing
(SE): Spherical equivalent
(OR): Odds ratio
(AL): Axial length
(SNPs): Single-nucleotide polymorphisms
(AF): Allele frequency
(MAF): Minor allele frequence
(GQ): Genotype quality
(DP): Depth
(PC): Principal
(VEP): Variant Effect Predictor
(BWA): Burrows-Wheeler Aligner
(PTVs): Protein-truncating variants
(CADD): Combined Annotation Dependent Depletion
(SKAT): SNP-Set (Sequence) Kernel Association Test
(GLM): Generalized linear model
(scRNA-seq): Single-cell RNA-seq
(CHB): China Beijing
(CHS): China South
(CDX): China Dai
(JPT): Japan
(KHV): Korean
(1KG): Genome Project
(GVCF): Genomic variant call format
(D): Diopters
(OD): Oculus Dexter
(OS): Oculus Sinister

## Availability of data and materials

The data that support the findings of this study are available from the corresponding author upon reasonable request.

## Authors’ contributions

The study was conceived, designed, and supervised by Xinjie Mao. Fan Lu, Shilai Xing and Xiangyi Yu. Analysis of data was performed by Xiangyi Yu, Ruijing Xia, Zhenlin Du, Xiaoguang Yu and Lulu Peng. The manuscript was written by Xiangyi Yu and Ruijing Xia. Written informed consent for publication was obtained from all participants.

## Competing interests

The authors declare no competing financial interests.

## Ethics approval and consent to participate

The protocol and documentation conducted in this study received full approval from institutional ethics committee of Wenzhou Eye and Vision Hospital with approval number 2023-059-K-48-05. Written informed consent was obtained from each participant.

## Acknowledgements

We express our gratitude to all participants in this study.

## Consent for publication

Not applicable.

## Funding

Not applicable.

---

**Additional file 1. Standard quality controls of 60 WGS samples.** Boxplot of (a) sample mean depth, (b) sample mean genotype quality, (c) sample mean call rate for 60 samples. (d) Principal component analysis plot comparing 60 individuals with East Asian populations from the 1000 Genomes Project.

**Additional file 2. Burden test of different type of variant.** Burden analysis used logistic regression in (a) all type of variants (b) common variants and (c) rare variants between cases and controls.

## Notes

### Competing Interest Statement

The authors have declared no competing interest.

### Funding Statement

This study did not receive any funding

### Author Declarations

Ethics committee of Wenzhou Eye and Vision Hospital gave ethical approval for this work

