## Supplementary figures and images for "Associations between RetNet gene polymorphisms and efficacy of orthokeratology for myopia control : sample from a clinical retrospective study"

### Addditional File 1

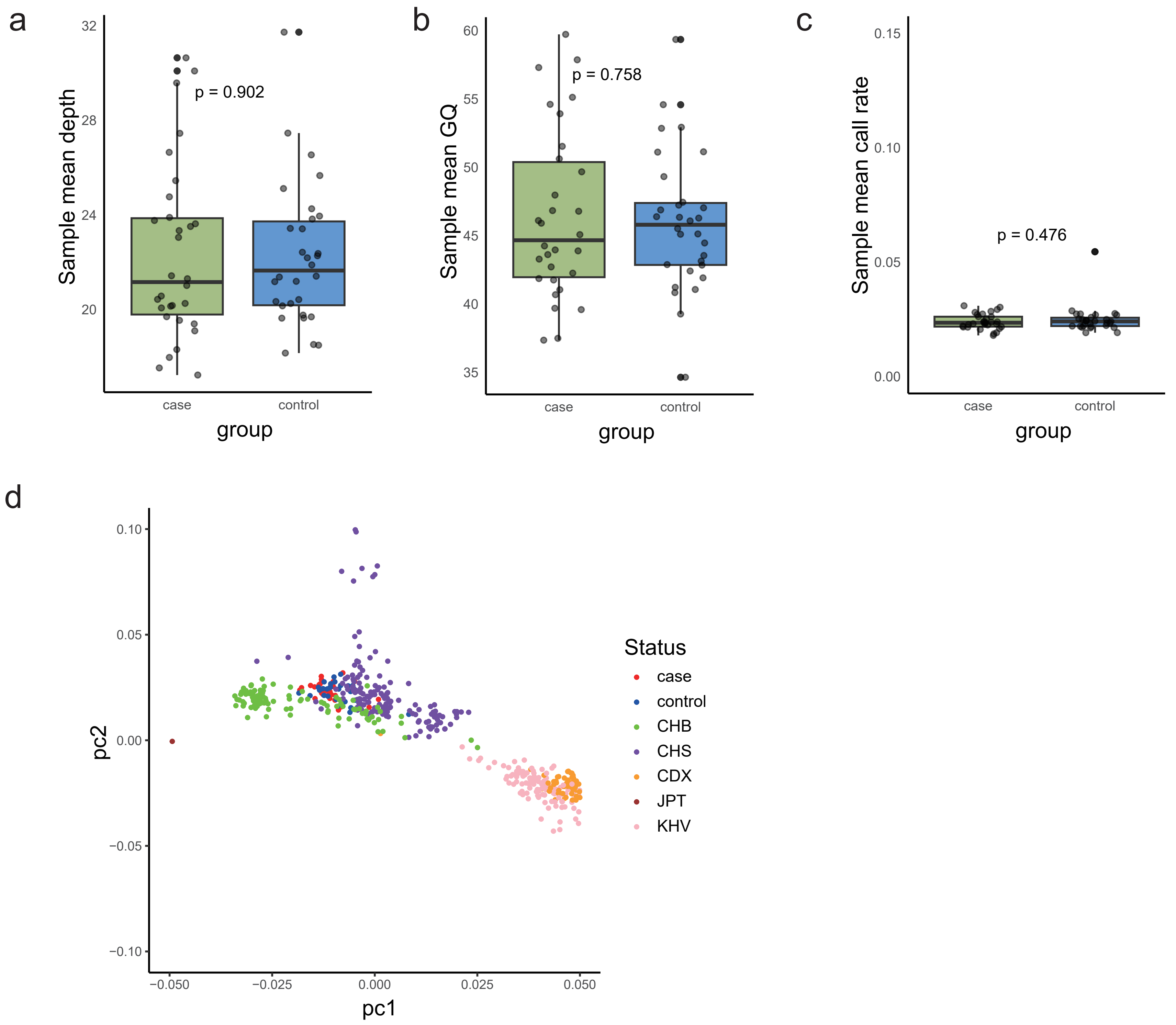

### Addditional File 2

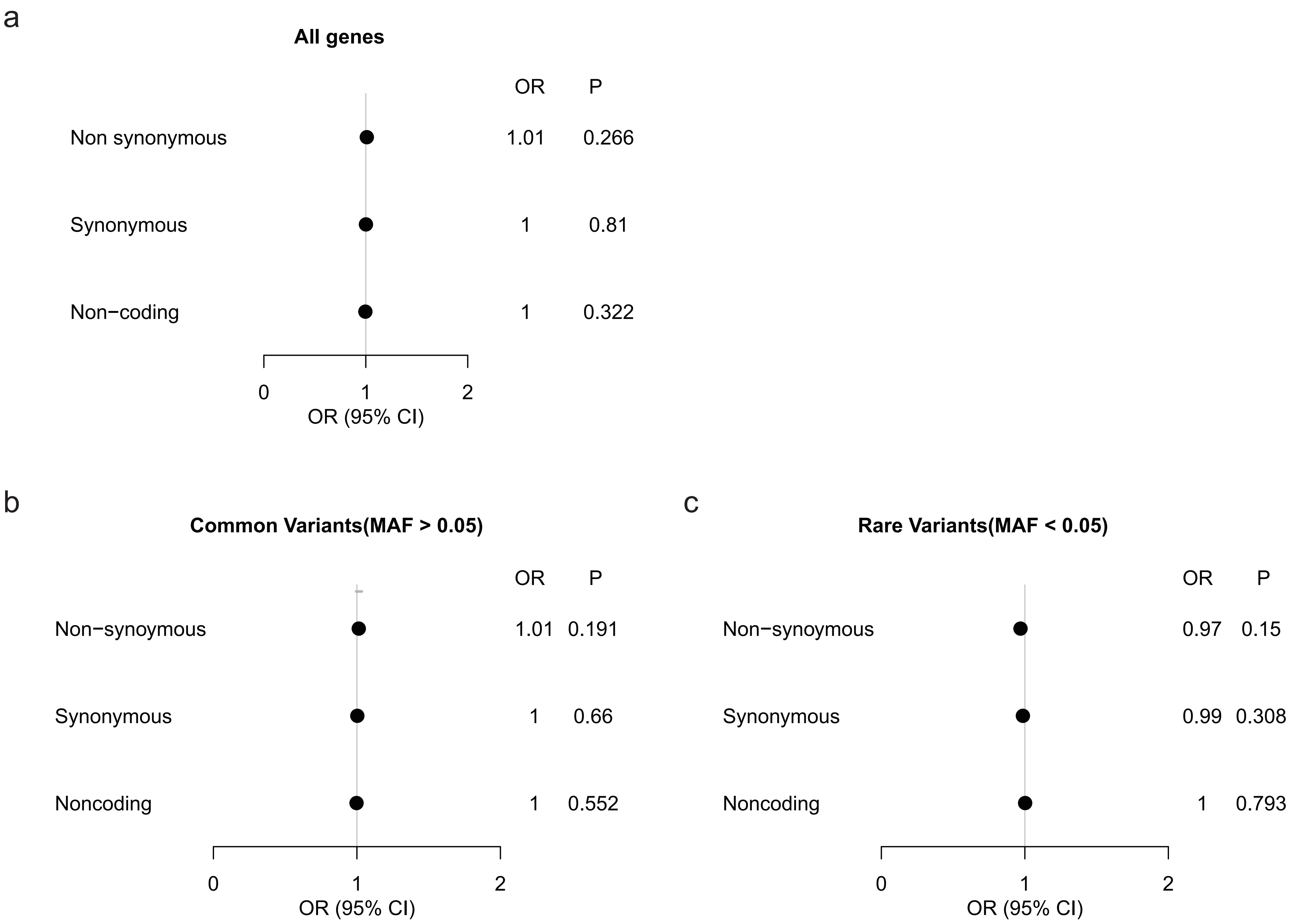
